# The RNA binding proteins ZFP36L1 and ZFP36L2 are dysregulated in airway epithelium in human and a murine model of asthma

**DOI:** 10.1101/2023.07.21.23293012

**Authors:** Jennifer Rynne, Elena Ortiz-Zapater, Dustin C. Bagley, George Doherty, Varsha Kanabar, David Jackson, Maddy Parsons, Jody Rosenblatt, Ian Adcock, Rocio T Martinez-Nunez

## Abstract

Asthma is the most common chronic inflammatory disease of the airways. The airway epithelium is a key driver of the disease, and numerous studies have established genome-wide differences in mRNA expression between health and asthma. However, the underlying molecular mechanisms for such differences remain poorly understood. We investigated the expression and possible role of the tristetraprolin (TTP) family of RNA binding proteins (RBPs), which are poorly understood in asthma. The human TTP family is comprised of *ZFP36*, *ZFP36L1* and *ZFP36L2,* and has essential roles in immune regulation by determining the stability and translation of myriad mRNAs encoding for inflammatory mediators. We found *ZFP36L1* and *ZFP36L2* mRNA levels significantly downregulated in the airway epithelium of patients with very severe asthma in different cohorts (5 healthy vs 8 severe asthma; 36 moderate asthma vs 37 severe asthma on inhaled steroids vs 26 severe asthma on oral corticoids). Integrating several datasets allowed us to infer that mRNAs potentially targeted by these RBPs are increased in severe asthma. *Zfp36l1* was downregulated in the lung of a mouse model of asthma, and immunostaining of *ex vivo* lung slices with a dual antibody demonstrated that Zfp36l1/l2 nuclear localization is increased in the airway epithelium of an acute asthma mouse model. Immunostaining of human bronchial biopsies showed that airway epithelial cell staining of ZFP36L1 was decreased in severe asthma as compared with mild, while ZFP36L2 was upregulated. We propose that the dysregulation of ZFP36L1/L2 levels as well as their subcellular mislocalization contributes to changes in mRNA expression and cytoplasmic fate in asthma.

## 1 Introduction

Asthma is a common chronic respiratory disease affecting between 1-29% of the population in different countries (1, 2, 3). It is characterised by variable symptoms of wheeze, chest tightness, shortness of breath and variable expiratory airflow limitation. Asthma is often defined as an inflammatory disease, implying that asthma has a major immune-related component. However, it is well established that structural cells including smooth muscle and airway epithelium play a major role in the disease.

Airway epithelial cells lie at the interface between the lung and the external environment, primarily acting as a protective barrier but also as immune modulators (4, 5). The importance of airway epithelial cells is well described in the pathophysiology of asthma. Patients with asthma present altered barrier function, mucus overproduction by goblet cells, epithelial cell damage and impaired repair. All these features contribute to airway remodelling, which is a broad term to define the airway structural changes that are always present in asthma (6). Multiple genome-wide approaches have been implemented to further our understanding of asthma and the role and profile of the bronchial epithelium in patients with this disease. These include RNA expression analysis (7, 8), breathomics (9), metabolomics (10) or sputum proteomics (11). However, there is poor insight into the underlying mechanisms regulating gene expression or driving their phenotype at the molecular level.

From transcription to translation into protein, RNA undergoes multiple steps such as splicing, transport and stability, encompassed under ‘post transcriptional regulation’. Post transcriptional regulation is mainly undertaken by microRNAs and RNA binding proteins (RBPs). Most omics approaches overlook these regulatory mechanisms and consider that mRNA expression is synonymous with corresponding protein levels. However, not all RNAs encode for proteins and there is little mRNA-to-protein correlation in many coding genes (12, 13). To further our understanding of mRNA cytoplasmic fate, we developed subcellular fractionation and RNA-sequencing (Frac-seq) (14). Frac-seq enables analysing mRNA steady levels (transcriptional) and those of transcripts bound to the translation machinery (ribosomes), the latter a better proxy for protein levels (15). Our previous work showed the disconnection between steady and ribosome-bound mRNA levels in bronchial epithelial cells from patients with asthma (16). We identified a network of six microRNAs that accounted for roughly 50% of the changes we observed in mRNA dysregulation (16). Thus, the remaining 50% levels must be mostly driven by RBPs. There is some evidence of RBP levels being dysregulated in airway epithelium in other respiratory diseases such as chronic obstructive pulmonary disease (17), however, their role in airway epithelium in asthma remains poorly understood.

The tristetraprolin (TTP) family of RBPs has been implicated in the regulation of immune responses (18), but little is known about their role in epithelium or asthma. These RBPs inhibit mRNA expression by binding to AU-rich elements (ARE) present in the 3’UnTranslated Region (UTR) of their target RNAs (19, 20). TTP is known to modulate the effect of glucocorticoids (21), and we have recently discovered that ZFP36L1/L2 modulate the effect of glucocorticoids and the expression levels of mRNAs encoding epithelial-related functions (22). Considering that glucocorticoids are the mainstay treatment of patients with asthma (23), we hypothesised that the TTP family may be dysregulated in asthma and set out to investigate their expression and potential roles in airway epithelium of human samples and asthma murine models.

## 2 Results

### 2.1 ZFP36L1 and ZFP36L2 mRNA levels are down regulated in bronchial epithelial cells from patients with severe asthma

We initially mined our previous Frac-seq dataset to determine the levels of all TTP members. Patient details were reporter previously (16). Changes in mRNA levels were determined by comparing total (Total) and polyribosome bound (Polyribosome) mRNA in health vs asthma. Polyribosome fractions excluded the monosomal (80S or one ribosome) fraction. We observed significantly decreased binding of *ZFP36L1* and *ZFP36L2* mRNAs to polyribosomes in primary bronchial epithelial cells from severe asthma patients compared to age- and sex- matched healthy controls (both p < 0.05, Figure 1A). Comparing total mRNA from healthy controls to severe asthma patients showed no difference in the expression of *ZFP36*, *ZFP36L1* or *ZFP36L2* mRNA (Figure 1A).

**Figure 1.**
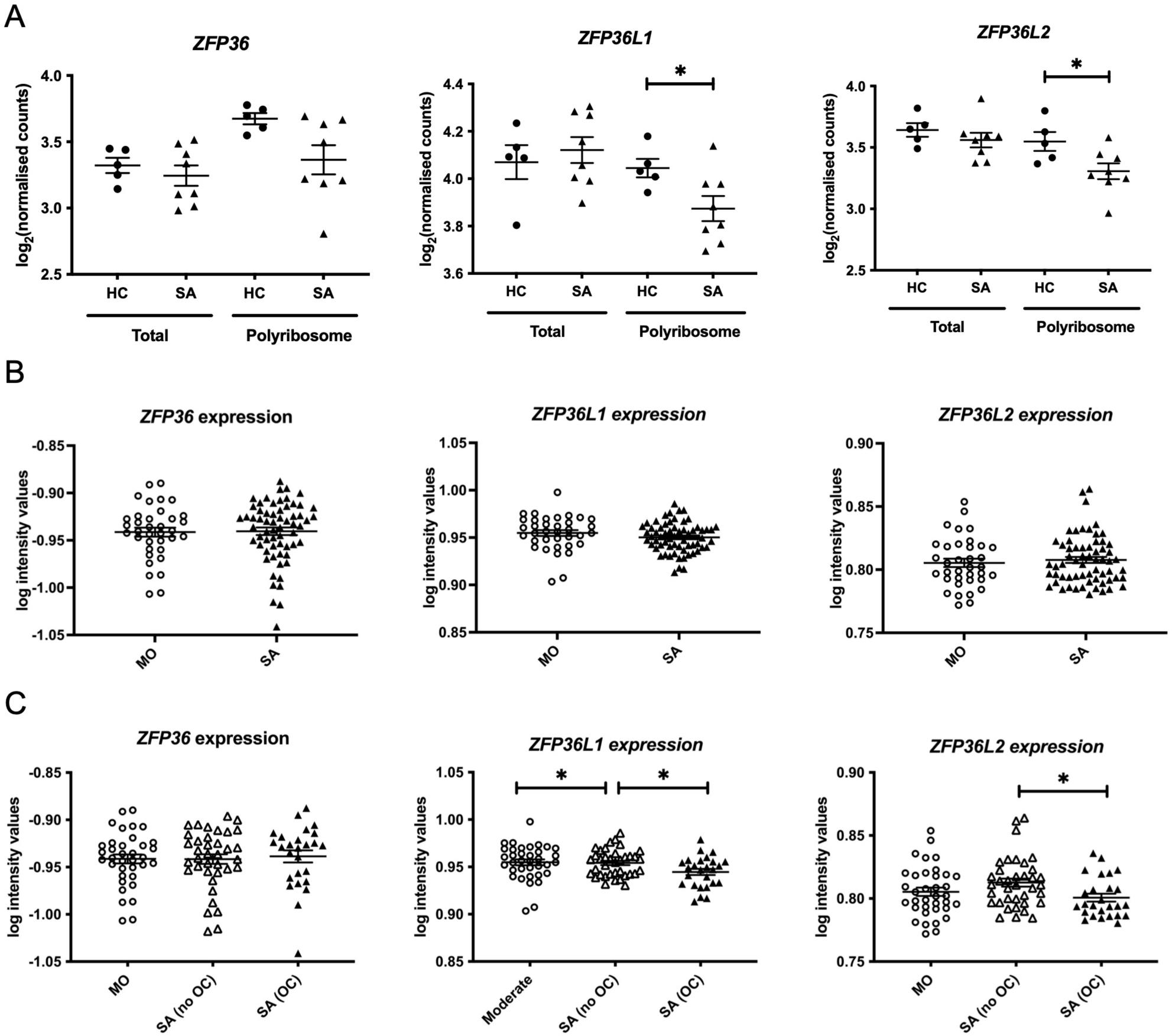
Analysis of the expression of the TTP family in human bronchial epithelial samples. (**A**) Frac-seq data showing the log counts for *ZFP36*, *ZFP36L1* and *ZFP36L2* in bronchial epithelial cells from severe asthma (SA, n=8) vs healthy control (HC, n=5) donors from (16) in total mRNA (Total) or polyribosome-bound mRNA (Polyribosome). (**B)** Analysis of GSE76227 dataset from U-BIOPRED showing the log intensity values of microarrays in bronchial brushings from patients with moderate (MO) and severe asthma (SA). (**C**) Analysis of GSE76227 dataset from U-BIOPRED showing the log intensity values of microarrays in bronchial brushing from patients with moderate (MO, n=36), severe asthma on oral corticoids (SA (OC), n=26) and severe asthma on inhaled GCs (SA(no OC)), n=37). Statistical significance was assessed by two-tailed t-tests on log transformed data. * p < 0.05.

These data prompted us to further interrogate the levels and role of these RBPs in asthma in a larger cohort. We analysed datasets from U-BIOPRED, the largest European consortium of severe asthma patients. The U-BIOPRED dataset (GSE76227) consists of transcriptomic arrays from bronchial brushings, which are enriched in bronchial epithelium (24). The authors compared moderate asthma patients on inhaled glucocorticoids (MO), representing patients with disease control, to severe asthma patients (SA), without differentiating between corticosteroid use. SA patients taking oral corticosteroids represent patients with inadequate control of disease. Figure 1B depicts graphs displaying the log intensity values from the microarray probes showing that there was no difference in *ZFP36, ZFP36L1* or *ZFP36L2* mRNA expression. Upon stratification of severe asthma patients into those on inhaled glucocorticoids (SA-no OC) or taking oral glucocorticoids (SA-OC), we observed downregulation of *ZFP36L1* mRNA expression in SA-no OC patients compared to MO patients (p < 0.05), and a further downregulation in SA-OC versus SA-no OC patients (p < 0.05, Figure 1C). We also found that the expression of *ZFP36L2* mRNA was downregulated in SA-OC patients compared to SA-no OC (Figure 1C). We did not observe differential expression of *ZFP36* mRNA levels.

### 2.2 ZFP36L1 and ZFP36L2 modulate genome-wide expression changes in bronchial epithelium in asthma

We further explored the potential role of ZFP36L1 and ZFP36L2 in bronchial epithelial cells in asthma. Our Frac-seq dataset consisted of mainly basal epithelial cells, while bronchial brushings from U-BIOPRED will contain a mixture of different airway epithelial cells. We interrogated the Lung Cell Atlas datasets comparing very mild patients with asthma not on inhaled corticosteroid therapy vs healthy controls (25). In health, *ZFP36L1* mRNA was expressed quite broadly in most epithelial cell types while *ZFP36L2* mRNAs appeared more present in Basal 1 and Basal 2 cells (Supplementary Figure 1). In asthma samples, *ZFP36L1* mRNA was decreased in mucous ciliated cells while it appeared to increase in ionocytes, ciliated and basal cycling cells (Figures 2A and 2B). *ZFP36L2* mRNA showed a trend towards downregulation in most airway cell types, with only submucosal and Club cells showing an increase in expression. Thus, single cell data support the concept that RBP mRNA expression is decreased in specific epithelial cell subsets in asthma.

**Figure 2.**
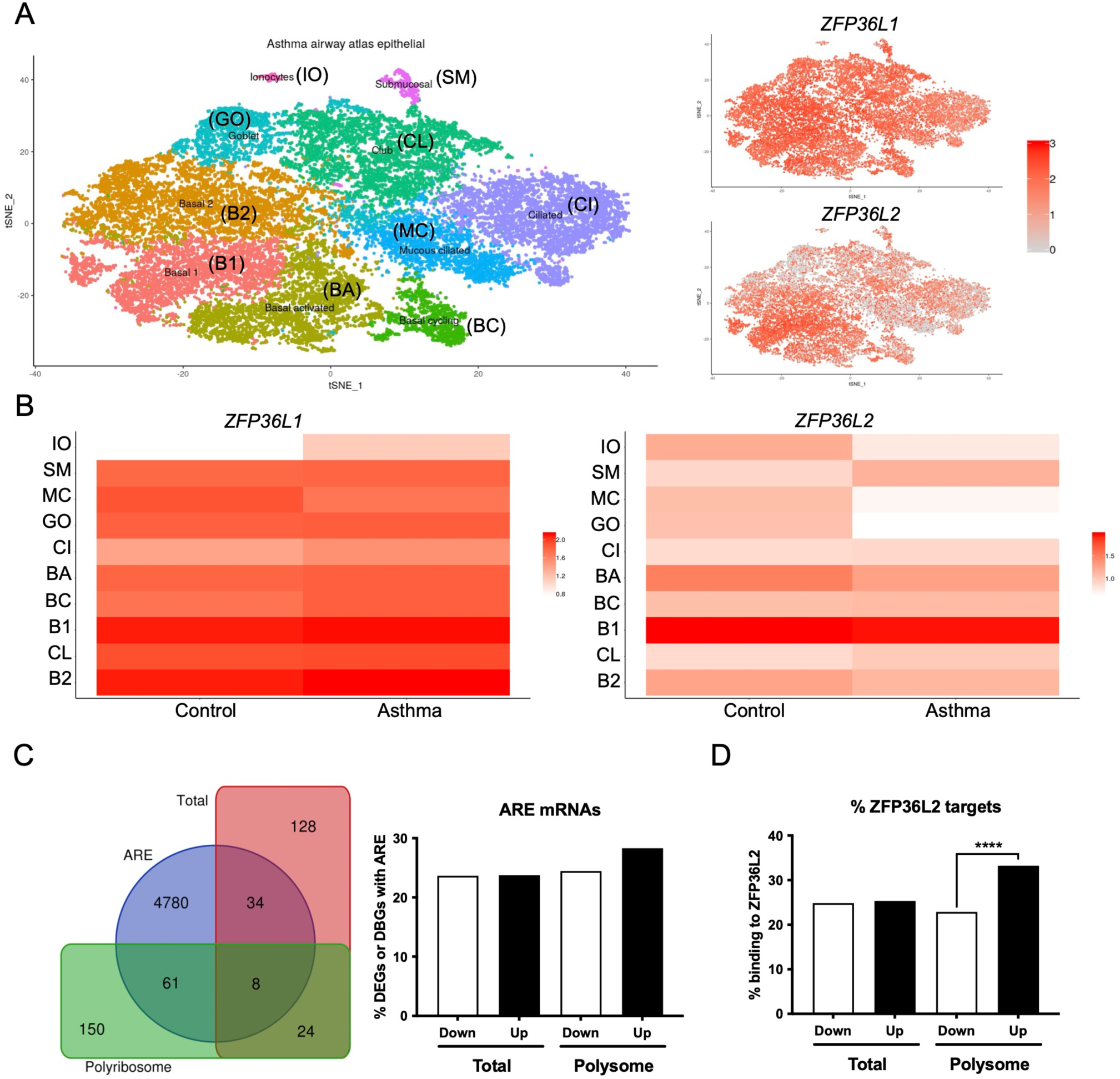
Airway epithelial cell-specific patterns of *ZFP36L1* and *ZFP36L2* mRNA expression and ZFP36L2 targets’ behaviour in severe asthma. (**A**) t-Distributed Stochastic Neighbor Embedding (t-SNE) plots representing the different airway epithelial cell types present in very mild asthma as per (25) (left), with *ZFP36L1* and *ZFP36L2* mRNA expression patterns represented on the tSNE’s on the right. (**B**) Relative expression *ZFP36L1* and *ZFP36L2* mRNA levels between health and asthma per airway epithelial cell type. IO: Ionocytes, SM: Submucosal, MC: Mucous ciliated, GO: Goblet, CI: Ciliated, BA: Basal activated, BC: Basal Cycling, B1: Basal 1, CL: Club, B2: Basal 2. (**C**) Left: Venn diagram representing the overlap of differentially expressed genes (DEGs in Total, red) or differentially bound genes (DBGs in Polyribosome, green) containing AU-rich element (ARE, blue) (26) when comparing health vs severe asthma. Right: bar plot showing the proportion and direction of changes (Up or Down regulated) of ARE-containing mRNAs in DEGs and DBGs when comparing health vs severe asthma. (**D**) Bar plot showing the proportion of ZFP36L2 targets as per (27) present per subcellular fraction in our Frac-seq health vs severe asthma dataset. Statistics were done employing a two-sided Chi-square test. ****: p < 0.0001.

We next analyzed if mRNAs containing AREs (26) were dysregulated in our Frac-seq dataset comparing health vs severe asthma (16). ARE-containing transcripts are targets of ZFP36L1 and ZFP36L2. Although we observed that a proportion of differentially expressed genes (DEGs) in total mRNA and differentially bound genes to polyribosome (DBGs) contain ARE in their 3’UTR (Figure 2C), these did not appear to be significantly enriched amongst DEGs or DBGs. To determine if ZFP36L2, specifically, may target DEGs and DBGs, we extracted known targets to be bound by ZFP36L2 from (27) and analyzed their presence in our Frac-seq DEGs and DBGs. Figure 2D shows that direct targets of ZFP36L2 were predominantly upregulated in polyribosome-bound genes in severe asthma. This enrichment was absent in total DEGs between healthy controls and severe asthma patients.

Considering that patients with severe asthma are treated with high doses of inhaled and/or oral corticosteroids, we also investigated if the changes seen in the DEGs and DBGs between health and SA were due to glucocorticoid (GC) exposure. We cross referenced our health vs severe asthma Frac-seq dataset with our recent Frac-seq data investigating post-transcriptional changes induced by GCs in primary bronchial epithelial cells (22). To that end, we compared upregulated and downregulated DEGs and DBGs between health and severe asthma and upregulated and downregulated DEGs and DBGs upon GC exposure. We only found 5 DEGs and 1 DBG between healthy controls and severe asthma that followed the same trend (up- or down-regulated in a fraction-dependent manner) upon GC treatment). We also investigated the expression of these mRNAs in the U-BIOPRED dataset and found only 3 were present, namely *DUSP1*, *SRM*, and *PTK2B*. These genes were not altered between moderate and severe asthma patients, or between severe asthma patients stratified by OCS usage (Supplementary Figure 2). These data suggest that decreased RBP expression in very severe asthma is unlikely to be due simply to oral corticosteroid use.

In summary, our analysis inferred that *ZFP36L1* and *ZFP36L2* encoding mRNAs are present in distinct airway epithelial cells and that ZFP36L2 preferentially targets mRNAs that are bound to polyribosomes in bronchial epithelial cells from patients with severe asthma.

### 2.3 Zfp36l1 mRNA levels are downregulated in the lungs of mice with asthma-like characteristics

After having analysed the levels of ZFP36L1 and ZFP36L2 in omics mRNA datasets and inferring their role in ARE regulation and epithelial cell biology, we interrogated their expression levels in an *in vivo* model of asthma. The House Dust Mite (HDM) model is an established model of allergen-induced inflammation and one of the most used in vivo models of asthma (28). C57BL/6J mice were administered intranasally 25 □l (1 mg/mL) protein weight solution dissolved in PBS of HDM extract (Citeq) or equal volumes of PBS five times a week for one week or five weeks, to produce and acute or chronic inflammation. This model has already been published elsewhere (29, 30) but we tested the effect of the HDM at different times using both H&E staining from FFPE sections of mice from the different groups (Figure 3A). We can observe immune infiltration in the section of HDM mice, increased after chronic exposure of HDM. After 5 weeks, the increase in the epithelium height is also evident (see arrows in Figure 3A). Total RNA from whole lung tissue from the different four groups was also analysed for the expression of two asthma-associated inflammatory cytokines, *Ccl20* and *Il13*. As expected, there was a clear increase in the expression of both transcripts after 1 week of HDM treatment (Figure 3B) and 5 weeks of HDM treatment (Figure 3C). We then assessed the mRNA levels of both *Zfp36l1* and *Zfp36l2* after both acute and chronic HDM treatment. As shown in Figure 3D, the expression of both *Zfp36l1* and *Zfp36l2* was not modified between PBS and acutely HDM treated mice. However, the mRNA expression of *Zfp36l1* in total lung tissue was significantly downregulated after chronic HDM exposure (Figure 3E, left panel). This was not seen for *Zfp36l2*, although there was lower expression compared to PBS mice in around half of the mice (Fig 3E, right panel).

**Figure 3.**
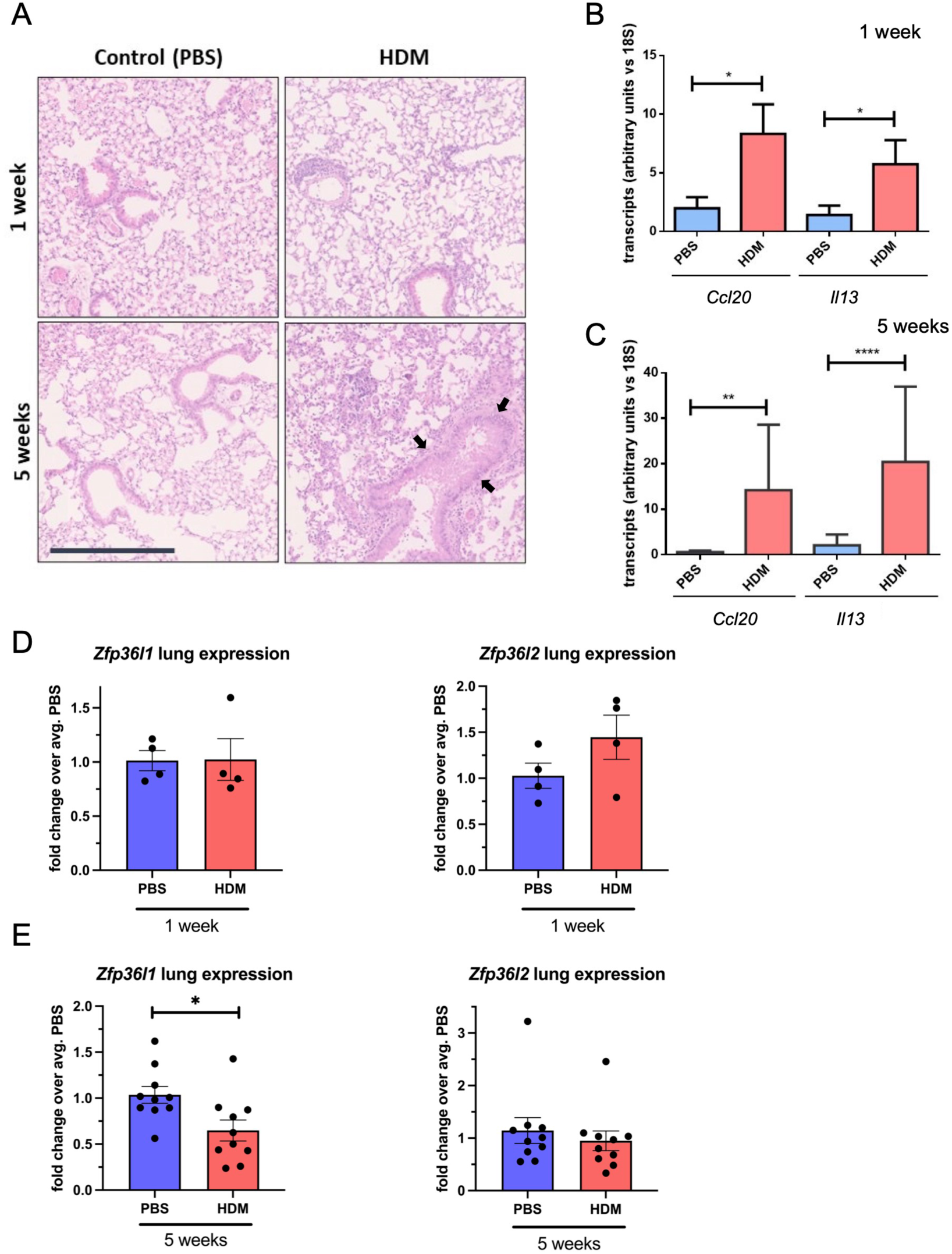
*Zfp36l1* and *Zfp36l2* mRNA expression in the lungs of mice with different types of asthma. (**A**) Representative images of Hematoxilyn and Eosin (H&E) staining of FFPE lung sections from mice treated with PBS as a control or with HDM for different times as indicated in the figure. Scale bars 125 μm. (**B**). The expression of two cytokines (*Ccl20* and *Il13*) in whole lung mRNA was analyzed by quantitative qPCR for 1 week and in (**C**) for 5 weeks. Bar graphs represent mean fold change (±SEM) over the average PBS-treated control (n=3 mice per group at PBS/HDM for 1 week and n=5 mice per group in 3 independent experiments for the 5 weeks treatment). (**D**) Expression of both *Zfp36l1* and *Zfp26l2* in whole lung mRNA analysed by quantitative PCR. Statistical significance was assessed by multiple two-tailed parametric t-tests on log transformed data. p < 0.05*, p<0.01** and p<0.001***.

### 2.4 Zfp36l1/l2 are mis-localized in the airways of mice with asthma-like lung characteristics

To further investigate our findings *in vivo*, we assessed the localization of Zfp36l1 and Zfp36l2 in the airways of HDM exposed mice. We employed mice exposed to HDM over three weeks; these mice produced a robust T2-driven inflammatory response and asthma-like phenotype including airway hyperresponsiveness, mucus hyperproduction, and immune cell infiltration. PBS- and HDM-treated mice were sacrificed and lungs harvested and processed to obtain precision-cut *ex vivo* lung slices (PCLS) that were fixed and analyzed by immunofluorescence and confocal imaging. Indeed, in healthy airways and bronchioles there was little to no Muc5ac (a pathological mucin highly upregulated in asthma) expression in the epithelial monolayer, nor immune cell infiltrate near airways in the surrounding alveolar space (Figure 4A, left). Unsurprisingly, HDM-treated airways had drastically increased expression of Muc5ac in the epithelium and significant immune cell infiltration near airways as shown by Ly-6G+ neutrophils (immune cell type notoriously increased in many asthmatics). We found that Zfp36l1/Zfp36l2 (using a dual-staining antibody for both Zfp36l1 and Zfp36l2 proteins) were expressed in airway epithelial cells and nearly evenly distributed between the nucleus and cytoplasm in healthy mice. However, in airways exposed to HDM, Zfp36l1/Zfp36l2 became robustly recruited into the nucleus of epithelial cells (Figure 4C). To confirm this, measurements of fluorescence intensity were made for nuclear and cytoplasmic Zfp36l1/Zfp36l2 and ratios calculated, demonstrating a highly significant increase in the nuclear signal of Zfp36l1/Zfp36l2 *ex vivo* in the airways.

**Figure 4.**
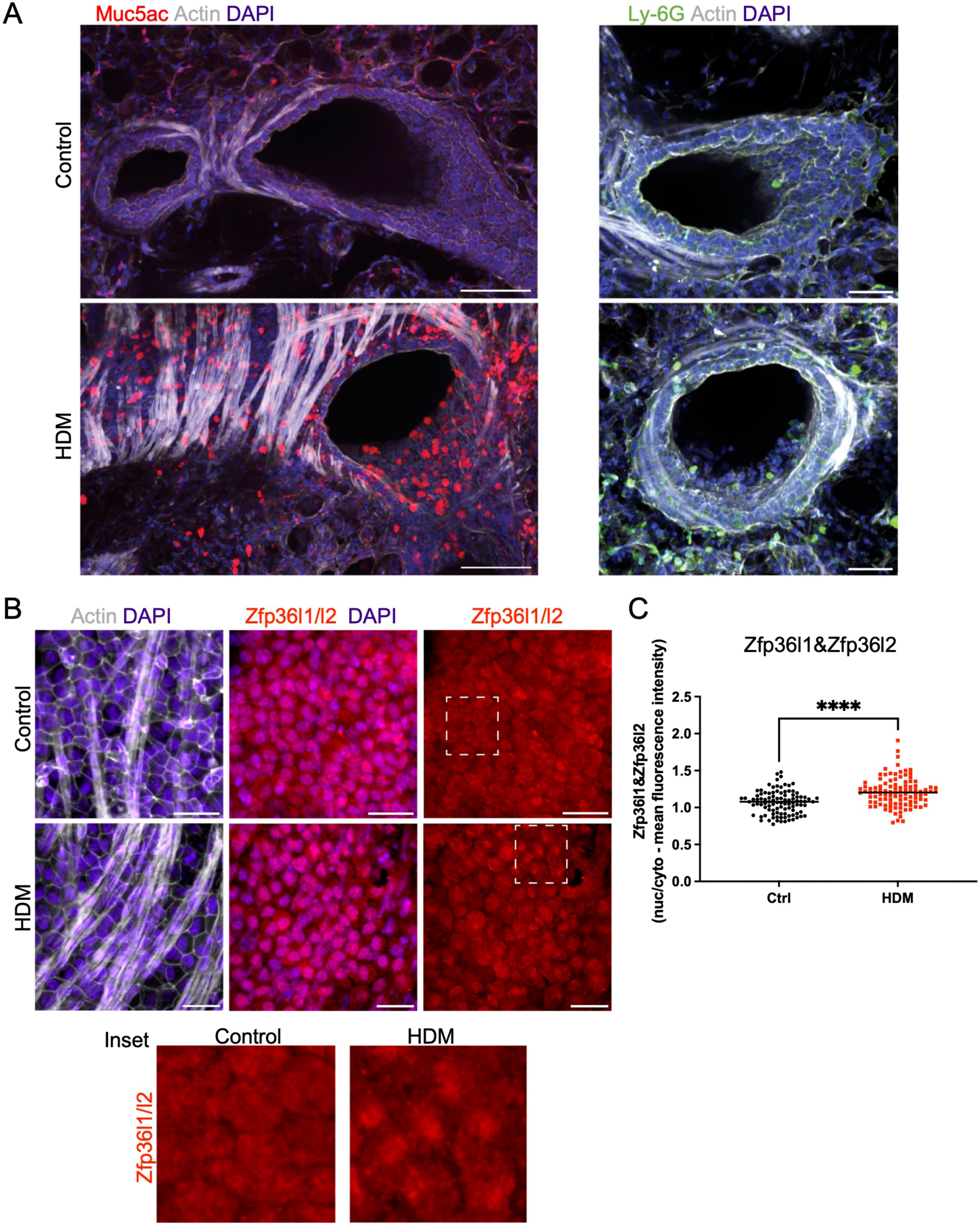
Intracellular mislocalization of Zfp36l1/Zfp36l2 in airway epithelial cells of mice with asthma-like characteristics. (**A**) Representative spinning disc confocal images of *ex vivo* lung slices from healthy controls and HDM-treated (three weeks) mice were fixed and immune-stained for asthmatic markers of mucus (Muc5ac) and neutrophils (Ly-6G) to demonstrate an asthma phenotype (scale bars are 100 and 50 microns, respectively). (**B**) Confocal projections of healthy and HDM-treated airway epithelial cells immune-stained for BRF1/2, actin and nuclei (scale bar 50 microns), with insets highlighting Zfp36l1/Zfp36l2 intracellular localization in epithelial cells in health (Control) and asthma (HDM). (**C**) Quantification of nuclear and cytoplasmic mean fluorescence intensity of Zfp36l1/Zfp36l2 in control and HDM-treated airways, using regions of interest (ROIs) defined by the DAPI (nuclei) and actin (cytoplasm) channels. Mann-Whitney test was performed on 100 cells from 3 mice per group. ****: p <0.0001. Ctrl: Control. HDM: House dust mite.

### 2.5 ZFP36L1 and ZFP36L2 immunostaining of bronchial biopsies differs with severity in human asthma

To further investigate the dysregulation of both *ZFP36L1* and *ZFP36L2* mRNA levels human (Figure 1) and mouse asthma (Figures 3 and 4) we performed immunostaining of ZFP36L1 and ZFP36L2 in bronchial biopsies from patients with different asthma severities and non-asthma controls. Our results show that immunostaining with both ZFP36L1 and ZFP36L2 was predominant in bronchial epithelium (Figures 5A and 5B) while also present in other cell types. Analysis of the positively stained airway epithelial cells showed that patients with mild asthma had increased staining of ZFP36L1 in epithelial cells as compared with healthy controls and severe asthma (Figure 5C). Patients with severe asthma showed low ZFP36L1 staining, significantly decreased as compared with mild but not healthy controls. ZFP36L2 staining was increased in patients with severe asthma as compared to both healthy controls and mild patients (Figure 5D), but there was no difference in the ZFP36L2 staining between healthy controls and mild asthmatic patients.

**Figure 5.**
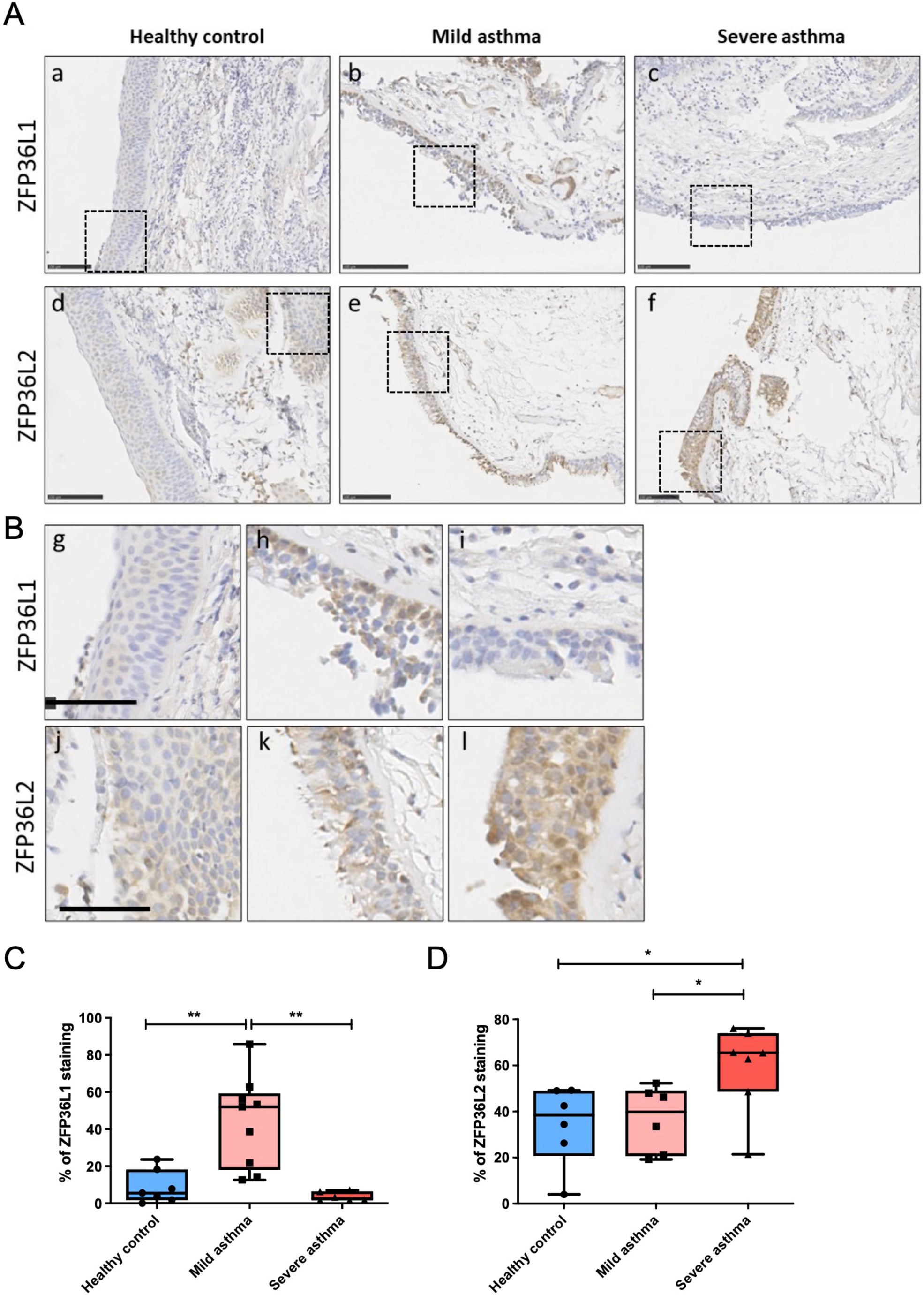
Immunohistochemistry staining of ZFP36L1 and ZFP36L2 in from bronchial biopsies from healthy controls and patients with mild and severe asthma. (**A**) Representative images of sections from patients classified as healthy controls (a and d), mild asthmatics (b and e) or patients with severe asthma (c and f). Stained was performed for ZFP36L1 and ZFP36L2 as indicated in the figure. Scale bar represents 100 μm. (**B**) Insets of the different sections shown in A. The scale bar represents 50 μm. (**C**) and (**D**). Quantification of ZFP36L1 and ZFP36L2 staining in FFPE sections from each experimental group. In **C**: Healthy controls n=7, Mild asthmatics n=9 and severe asthmatics n=6. In **D**, Healthy controls n=6, mild asthmatics n=6 and severe asthmatics n=7. Statistical significance was assessed by multiple two-tailed t-tests on log-transformed data. *: p < 0.05, **: p < 0.01.

Taken together, our data show that ZFP36L1 and ZFP36L2 are expressed in airway epithelial cells and that their levels and subcellular localization are modified in asthma, in a severity-dependent manner. These proteins alter post-transcriptional gene expression in bronchial epithelial cells in severe asthma, granting further investigation into their role in chronic airway inflammation.

## 3 Discussion

Our data demonstrate that ZFP36L1 and ZFP36L2 levels and subcellular localization are dysregulated in primary samples from both human and murine-like asthma. We have also performed *in silico* analysis of publicly available datasets of ZFP36L2 targets and observed that these are particularly enriched in genes that present increased binding to polyribosomes in severe asthma human primary bronchial epithelium. Together, our data strongly suggest that ZFP36L1 and ZFP36L2 drive changes in gene expression in primary epithelial cells in asthma.

Airway epithelial cells contribute to disease pathobiology in asthma through immune modulation, defective barrier function, and airway remodelling. All asthma phenotypes present epithelial cell damage (5, 31, 32, 33), and airway structural changes can be present before the onset of inflammation (34, 35). These observations are contrary to the old paradigm of repeated inflammation driving epithelial damage and deficient repair resulting in airway remodelling (36). Thus, epithelial changes appear to occur early on and be central to disease progression. It is possible that early epithelial cell gene expression reprogramming occurs leading to these cells behaving differently in asthma. This is indeed supported by studies that have determined a differential epigenetic signature in primary airway epithelium in asthma (37). It is also possible that relatively small changes in master-regulators, such as RBPs, cause a sustained effect that leads to a different profile and cellular behaviour.

RBPs can modulate mRNA transcript levels and fate by binding to their targets and controlling most post-transcriptional steps, including alternative splicing, export, stability, storage, decay or translation into protein. The role of post-transcriptional gene expression regulation in inflammation is well established (18, 38) and defective RNA-RBP interactions can contribute to dysregulated immune responses (39). Additionally, RBPs have been found dysregulated in the epithelium of patients with chronic obstructive pulmonary disease (17). There is no literature, to our knowledge, about the role of RBPs in asthma, where research in post-transcriptional regulation has focused on microRNAs (40).

Amongst RBPs, the TTP family has well-established roles in modulating the immune system (41, 42, 43), as well as the response to glucocorticoids (21, 22). Our data in Figure 1 showed a decreased expression of *ZFP36L1* and *ZFP36L2* mRNA in airway epithelial cells from patients with severe asthma, particularly of those that are undergoing oral corticosteroid treatment. We also found decreased of *Zfp36l1* and *Zfp36l2* mRNA in chronic asthma-like inflammation (akin to human severe asthma) in mice (Figure 3). At the protein level, in human bronchial biopsies (Figure 5), ZFP36L1 showed an increase in expression in mild patients, i.e. not taking glucocorticoids, as compared to healthy controls, and a decrease in patients with severe asthma (Figure 5). Contrary to mRNA expression, ZFP36L2 protein appeared upregulated in patients with severe asthma, i.e. on high doses of inhaled and/or oral glucocorticoids. These data highlight the importance of performing validations at the protein level, and the poor correlation existing between mRNA and protein levels (44). It is also important to highlight that immunohistochemistry is a good methodology to indicate where a protein is expressed, but it is semi-quantitative. Further analysis of protein levels in specific airway epithelial types employing other methodologies such as fluorescent western blotting could offer further insight into total protein levels.

Mechanistically, it possible that expression levels of ZFP36L1 and ZFP36L2 are driven by different mechanisms in, particularly, severe/chronic asthma. Our data on murine models of acute and chronic asthma-like inflammation (Figure 3) showed decreased of *Zfp36l1* mRNA in the lung of chronic asthma-like inflammation (akin to human severe asthma), although these data correspond to whole lung. More detailed time courses of HDM exposure and analysis of specific cell types may unravel the dynamics of expression of these RBPs. Zfp36l1/l2 nuclear localization was increased in the airways of mice with asthma-like characteristics (Figure 4). ZFP36L1 and predominantly ZFP36L2 are increased in bronchial epithelial cells exposed to glucocorticoids, while their nuclear localization is also enhanced upon glucocorticoid exposure (22). Noteworthy, our antibody staining for immunofluorescence (in both (22) and Figure 4 PCLS) did not allow us to distinguish between ZFP36L1/Zfp36l1 and ZFP36L2/Zfp36l2. In bronchial biopsies, we observed a trend towards increased nuclear localization for ZFP36L2 in patients with severe asthma (Supplementary Figure 3) while ZFP36L1 appeared more localised in the cytosol. Together, these data point towards chronic inflammation driving changes in ZFP36L1 expression in asthma, while glucocorticoids may be the main drivers of ZFP36L2 protein expression in severe asthma. In either case, it appears that there is an increase in nuclear localization for these proteins in asthma. While this may infer an increase in their nuclear role, our integration of ZFP36L2 targets showed that ZFP36L2 exerts its effects mainly by modulating polyribosome association of mRNAs in severe asthma (Figure 2C). Thus, it is possible that the changes exerted by ZFP36L2 in the cytosol are driven by is nuclear kidnapping.

ZFP36L1 and ZFP36L2 encoding mRNAs also contain ARE in their 3’UTRs and can therefore regulate the expression of one another. Although our ARE analysis did not show an enrichment in ARE-containing mRNAs in patients with severe asthma, ARE-containing mRNAs accounted for 20-28% of the genes differentially expressed and differentially bound to polyribosomes between health and severe asthma (Figure 2C). Further investigation into the specific dynamics of these two RBPs in primary epithelium will enable elucidating their potential influence on each other’s expression and/or targets. It is also possible that these proteins exert distinct effects in an airway cell-type manner. *ZFP36L1* and *ZFP36L2* mRNAs appeared differentially expressed in individual airway epithelial cell types, with these expression patterns varying between health and asthma (Figures 2A and 2B). This granularity is lost in bulk RNA-sequencing and these data suggest that these RBPs are indeed not redundant.

In summary, we provide, to our knowledge, the first evidence that RBPs are dysregulated in airway epithelial cells in human asthma and murine models of asthma in a severity-dependent manner. ZFP36L1/L2 appear to be modulated post-transcriptionally and post-translationally, with their subcellular localization influenced by chronic inflammation and possibly glucocorticoids. Recent evidence shows that ZFP36L1/L2 modulate epithelial-encoding mRNAs (22). We propose RBPs as novel modulators of inflammation and epithelial structure in asthma and their further investigation in chronic airway disease.

## 4 Materials and Methods

### 4.1 Human samples

Our Frac-seq cohort (n=5 healthy and n=8 severe asthma) and those utilized in biopsy staining were part of the Wessex severe asthma cohort (45), in which patients with severe asthma were defined as those fulfilling the European Respiratory Society/American Thoracic Society (ERS/ATS) criteria for severe asthma (23) and thus treated with high doses of inhaled and/or oral corticosteroids. Bronchial biopsies from Figure 5 were also part of the same cohort. We are using previously collected data that is publicly available. REC Numbers 06/Q0505/12 and 05/Q1702/165.

### 4.2 In vivo experiments: house dust mite (HDM) sensitization

All the research in this manuscript complies with ethical regulations. The use of animals for this study was approved by the Ethical Review Committee at King’s College London and the Home Office, UK. All animals were housed in the Biological Support Unit (BSU) located in New Hunt’s House at King’s College London. All experiments were carried out under project license number P9672569A and personal license number I0F9CA46A. For House Dust Mite (HDM) immune-sensitised protocol, 6-8 weeks female C57BL/6 mice, were anaesthetised with isofluorane and administered either 25Dg (total protein) of HDM extract (Citeq Biologics; 1 mg/ml protein weight solution dissolved in PBS) or 25Dl of PBS intranasally 5 times per week for 5 consecutive weeks. Control mice received 25Dl of PBS. Mice were culled 24 h after the final HDM or PBS dose. Mice were sacrificed in a CO2 gas chamber for lung dissection.

### 4.3 RNA extraction, reverse transcription and quantitative PCR (RT-qPCR)

RNA was extracted using TRIzol (Invitrogen) or TRIzol LS (Sigma-Aldrich) and reverse transcribed employing RNAse H Minus Reverse Transcriptase, Ribolock and random hexamers (ThermoFisher Scientific). qPCR was performed using NEB Luna buffer (New England Biolabs) and TaqMan assays (ThermoFisher Scientific).

### 4.4 Tissue processing and analysis of immunohistochemistry

Immunohistochemistry staining of ZFP36L1 and ZFP36L2 in lung tissue from bronchial biopsies in non-asthmatic patients, mild asthma, and severe asthma was carried out by Dr Jon Ward at the University of Southampton. Briefly, 10 µm thick sections paraffin-embedded sections were melted at 95°C for 2h and de-waxed by dipping slides in xylene 2 × 10 min, 100% EtOh 2 × 5 min, 70% EtOH 1×5min and 50% EtOH 1×5min. Antigen retrieval was carried using sodium citrate buffer (0.0874 M sodium citrate, 0.0126 M citric acid pH 6) and incubating the slides for 20 min in a pressure cooker at 95 °C. Endogenous peroxidase activity was blocked via 10 min incubation in hydrogen peroxide (3% in TBS) for DAB staining. Tissues were then washed 3× with TBS and non-specific binding was blocked via incubation with TBS-1%BSA-1%FBS blocking solution for 1 h at room temperature. Primary antibody was added to the tissues, and these were left at 4 °C overnight. After 3× washes with TBS, DAB staining was visualised by adding DAB developing solution for up to 20 min (Dako). Tissues were then counterstained using haematoxylin for 1 s. Finally, tissues were dehydrated with graded alcohols and xylene before being mounted with DPX. The slides were stained for ZFP36L1 (abx124297, Abbexa, 1:3000 dilution) and for ZFP36L2 (PA5-30644, Invitrogen, 1:1500 dilution). Immunohistochemistry analysis was done employing Qupath (https://qupath.github.io/) for quantitative analysis. Positive epithelial cells were counted and expressed as a percentage of the total epithelium per slide. Mild asthma patients had controlled asthma, while severe asthma samples were obtained from the Wessex Severe Asthma Cohort and were classified as having inadequately controlled disease and fulfilled the ERS/ATS criteria for severe asthma.

### 4.5 Precision-cut *ex vivo* lung slices (PCLSs)

Ex vivo lung slices were obtained from mice, within 48hrs of their last allergen priming challenge, adapted from the protocol of (46). Briefly, mice were humanely killed by CO2 inhalation followed by cervical dislocation. The lungs were inflated with 2% low melting agarose (Fisher – BP1360) prepared in HBSS+ (Gibco – 14025) before lungs, along with the heart and trachea, were excised, washed in PBS, and the lobes separated. Individual lobes were then embedded in 4% low melting agarose and solidified on ice. 200micron thick slices were cut on a Leica VT1200S vibratome and washed and incubated in DMEM/F-12 medium supplemented with 10% foetal bovine serum (FBS) and antibiotics.

### 4.6 Immunofluorescence and imaging of fixed PCLS

PFA fixed ex vivo lung slices were incubated for one hour at room temperature in blocking solution: PBS containing 0.1% triton X-100, 0.1% sodium azide, and 2% bovine albumin (BSA), before incubating overnight a 4°C at 1:100 in blocking solution for all primary antibodies used: mouse anti-Muc5ac (Abcam ab3649), rat anti-Ly-6G (Abcam ab2557), rabbit anti-BRF1/2 (Cell Signaling Technologies). Ex vivo lung slices were washed 3 × 30 minutes in PBS+0.5% Triton X-100) before incubating overnight at 4°C overnight with: 1:100 Alexa Fluor 488 goat anti-rabbit (Thermo Scientific - A11008) or anti-mouse (A32723) IgG, Alexa Fluor 568 goat anti-rabbit (A11011) or anti-mouse (A11004) IgG, or Alexa Fluor 647 goat anit-rabbit (A32733) or anti-mouse (A21235) IgG + 1:250 Alexa Fluor 488, 568, or 647 Phalloidin (Thermo Scientific – A12379, A12380, A22287, respectively). Slices were washed 3 × 30 minutes in PBS+0.5% Triton X-100, stained with 1:1000 DAPI in PBS for 20 minutes, mounted in ProLong Gold (Invitrogen P36930), and imaged on a Nikon Eclipse Ti2 spinning disc confocal microscope with a 20X or 40X objective.

### 4.7 Statistical analyses

GraphPad Prism and R studio software was used for the generation of graphs and analysis of data. All packages are available at CRAN. Two group analyses were done employing two-tailed tests. Enrichment analysis was performed using binomial tests. Differential proportions in Figure 2 were done employing a Chi-Square test. In all cases, p < 0.05 *, p < 0.01 **, p < 0.001 ***. RNA-seq analysis was evaluated using the DESeq2 package for differential gene expression, for details please see (22). Microarray analysis was performed using the limma package.

## 6 Author Contributions

JR EO-Z and DB acquired, analyzed and interpreted the data; GD and VK contributed experimentally; DJ, MP and JR contributed intellectually and/or financially; IA provided critical intellectual input; RTMN conceived and designed the work and analyzed data. All authors agreed to their inclusion in this work.

## 8 Acknowledgments

We thank all the participants of the Wessex Severe Asthma Cohort and Dr Jon Ward (Immunohistochemistry Unit, University of Southampton) for his work immunostaining human bronchial biopsies.

## 9 Supplementary Material

Supplementary Figures are provided.

## 10 Data Availability Statement

The datasets for this study can be found in the Gene Expression Omnibus https://www.ncbi.nlm.nih.gov/geo/ under accession numbers GSE213495 and GSE76227.

