## Supplemental Figures for "The RNA binding proteins ZFP36L1 and ZFP36L2 are dysregulated in airway epithelium in human and a murine model of asthma"

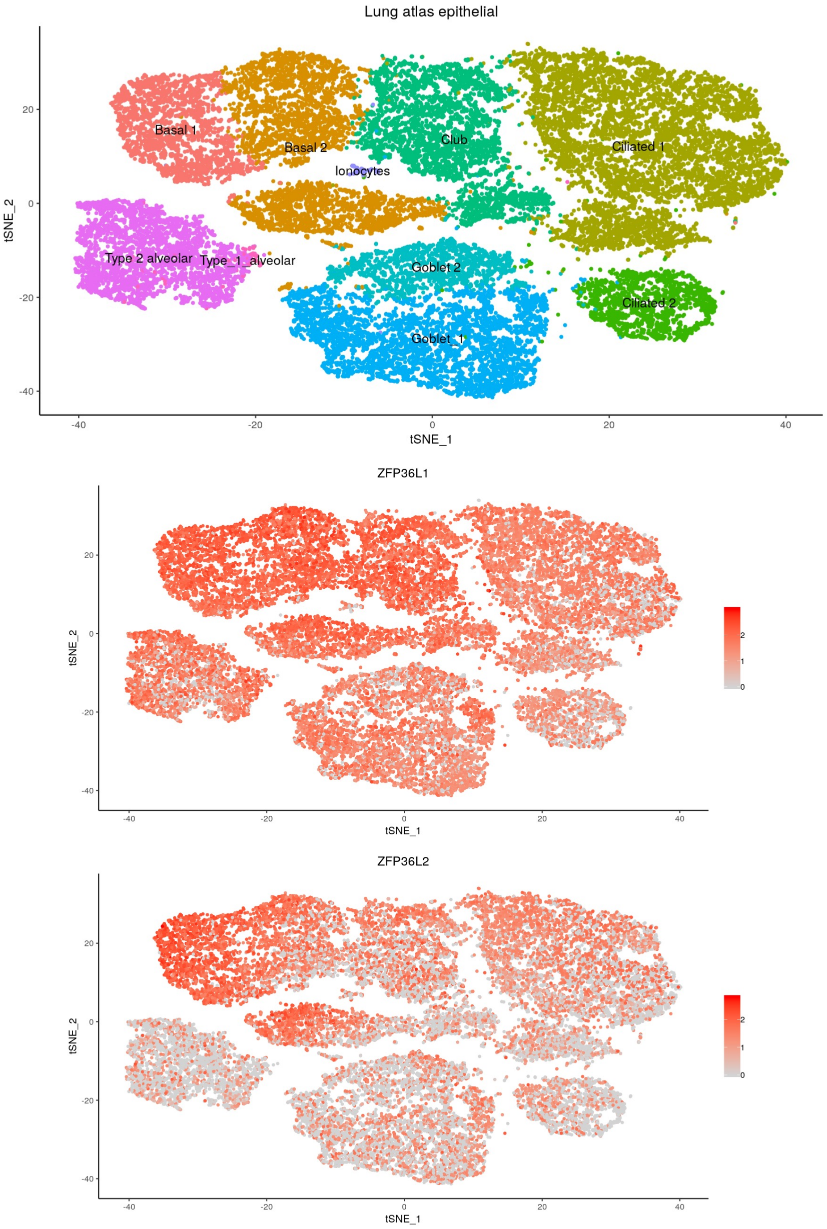


**Supplementary Figure 1. Epithelial cell expression of *ZFP36L1* and *ZFP36L2* encoding transcripts**. We interrogated the Lung Cell Atlas (https://asthma.cellgeni.sanger.ac.uk/) to determine the main airway epithelial cell type that presents ZFP36L1 and ZFP36L2 expression. While ZFP36L1 was more diffusely present, ZFP36L2 expression was more restricted to Basal 1 and Basal 2 type epithelial cells.

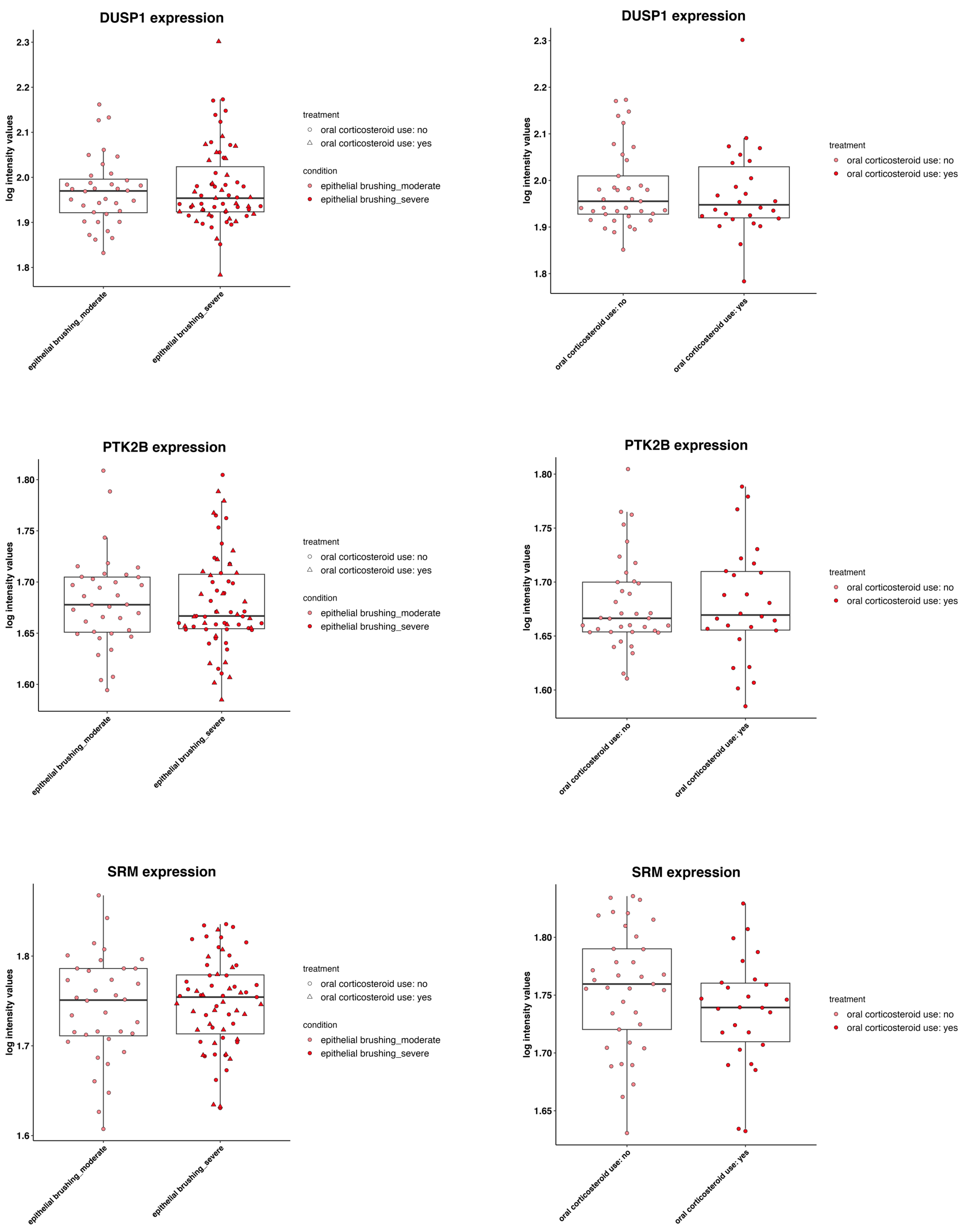


**Supplementary Figure 2. mRNA expression in asthma related transcripts in the U-BIOPRED dataset (GSE76227).**

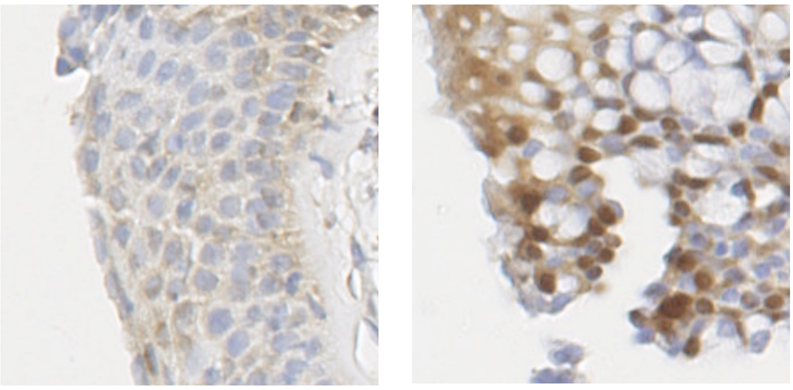

Healthy control                Severe asthma

**Supplementary Figure 3. mRNA expression in asthma related transcripts in the U-BIOPRED dataset (GSE76227).**
